# Duration of COVID-19 mRNA Vaccine Effectiveness against Severe Disease

**DOI:** 10.1101/2022.04.12.22273745

**Authors:** Devendra Bansal, Jazeel Abdulmajeed, Esraa Yassin, Maha H M A Al-Shamali, Soha S A Albayat, Sayed M Himatt, Farhan S Cyprian, Tawanda Chivese, Jesha M A Mundodan, Hayat Khogali, Rekayahouda Baaboura, Anvar H Kaleeckal, Mujeeb C Kandy, Ali Nizar Latif, Mohamed G Al-Kuwari, Hamad Eid Al-Romaihi, Abdullatif Al Khal, Roberto Bertollini, Mohamed H Al-Thani, Elmobashar Farag, Suhail A R Doi

**Author notes:** **Corresponding author** Suhail A. R. Doi MBBS, MMed, MClinEpid, PhD; FRCP(Edin), Professor of Clinical Epidemiology & Clinical Endocrinologist, Head, Department of Population Medicine, College of Medicine, Qatar University, P.O. box 2713, Doha, Qatar, E.

## Abstract

**Background:** Waning immunity following administration of mRNA based COVID-19 vaccines remains a concern for many health systems. We undertook a study of SARS-CoV-2 infections, with and without requirement for intensive care to shed more light on the duration of vaccine effectiveness for protection against the need for intensive care.

**Methods:** We used a matched case-control study design with the study base being all individuals with first infection with SARS-CoV-2 reported in the State of Qatar between 1 Jan 2021 and 20 Feb 2022. Cases were those requiring intensive care while controls were those who recovered without need for intensive care. Vaccine effectiveness against requiring intensive care and number needed to vaccinate (NNV) to prevent one more case of COVID-19 requiring intensive care were computed for the mRNA (BNT162b2 / mRNA-1273) vaccines.

**Results:** Vaccine effectiveness against requiring intensive care was 59% (95% confidence interval [CI], 50 to 76) between the first and second dose and strengthened to 89% (95% CI, 85 to 92) between the second dose and 4 months post the second dose in persons who received a primary course of the vaccine. There was no waning of vaccine effectiveness in the period from 4 to 12 months after the second dose.

**Conclusions:** This study demonstrates that vaccine effectiveness against requiring intensive care remains robust till at least 12 months after the second dose of mRNA based vaccines.

## INTRODUCTION

The Pfizer-BioNTech (BNT162b2) and Moderna (mRNA-1273) mRNA based vaccines are given in two doses scheduled three to four weeks apart. Evidence is still accruing regarding the duration of protection afforded by these two mRNA vaccines. Using antibody kinetics, it has been suggested that vaccine effectiveness declines to approximately 70% at 250 days post-vaccination^1^ but this ignores the presence of non-serologic components of the immune response. Recent studies have raised concern over a more rapid waning of both antibody titers and vaccine effectiveness against any infection over time, especially among older populations ^2–4^. However, the latter seems more of a problem with protection against any infection rather than severe disease since Pfizer-BioNTech have reported a gradual decline in efficacy from 96% between 7 days & 2 months to 84% between 4 to 6 months against any infection but reported efficacy was 97% for severe disease in this period ^5^.

Results of other studies concur with the Pfizer-BioNTech report for vaccine effectiveness against hospitalization and/or severe disease, ranging between 84–96%, up to 6 months following vaccination^4,6–8^. The potential for waning immunity against severe disease remains a concern for many health systems and thus there are varying recommendations on the timing of a third dose. However, decision making to date has been driven mainly by protection against any infection because there are concerns that effectiveness against severe disease may show a similar pattern^9^. but there is a paucity of data regarding waning of immunity against severe disease. We therefore undertook an evaluation of those diagnosed with COVID-19 in the State of Qatar, with and without requirement for intensive care, between 1 Jan 2021 and 20 Feb 2022 to shed more light on the duration of vaccine effectiveness for protection against the need for intensive care.

## METHODS

This was a matched case-control study with the study base being all individuals with first infection with SARS-CoV-2 reported in the State of Qatar between 1 Jan 2021 and 20 Feb 2022. Cases were those requiring intensive care while controls were those who recovered without need for intensive care. Approval and consent to participate were obtained (Ethics approval ERC-826-3-2020) and waiver of informed consent was given by the Health Research Governance Department at the Ministry of Public Health. All data were deidentified before sharing for analysis.

### Data Sources

Demographic information and clinical characteristics data were obtained from the Ministry of Public Health (MoPH), Qatar database. The national surveillance system (Surveillance & Vaccine Electronic System) at MoPH receives PCR confirmed case notification from Hamad Medical Corporation (HMC). The dates of the first and second dose of the two-dose vaccine schedule as well as date of a third dose, if administered (commenced in September 2021 in Qatar), were retrieved from the Primary Health Care Corporation (PHCC) and intensive care admissions data were obtained from HMC and these were linked to the MoPH database. These linked databases constituted the national federated databases for COVID-19 in Qatar.

### Vaccination

Majority of the population received one of the two mRNA vaccines as the primary two dose schedule three to four weeks apart although very few residents (1.6%) also received the AstraZeneca (ChAdOx1) vaccine as well, usually administered outside Qatar. The same brand of mRNA vaccine was used in the booster as in the primary series in the majority of the population. For the booster dose of mRNA-1273, half the dose used in the primary series was administered. Booster doses were initially administered 8 months after the second dose but later this was reduced to six months because of concerns regarding possible waning of protection from the primary schedule.

### Variables of interest

The MoPH database contains demographic and comorbidity information on all persons residing in Qatar who tested positive for SARS-CoV-2. The comorbidities included a range of chronic conditions (diabetes, hypertension, other cardiovascular diseases, asthma/COPD, cerebrovascular disease, rheumatological diseases, cancer, kidney disease, neurological disease, hematological disorders, immunity related disorders, liver disease and obesity) and this was classified as none, 1 to 4 and >4 conditions for this study. Infection in relation to each vaccine course was divided into seven intervals: 0 - infected when unvaccinated or infected prior to the first dose of the vaccine, 1 - infected in the period between the first and second dose, 2 - infected after the second dose and till four months after the second dose, 3 - infected four to six months after the second dose 4-infected six to nine months after the second dose, 5-infected nine to twelve months after the second dose and 6-infected after the third (booster) dose (data available from September 2021 to February 2022).

### COVID-19 Testing Data

PCR testing for SARS-CoV-2 in Qatar is undertaken by the HMC and details regarding the laboratory methods have previously been published^10^. More recently, rapid antigen tests were also introduced for testing at health care facilities on or after January 5, 2022. Swab collection is carried out at HMC, PHCC, other governmental, semi-governmental, and private health institutions across the country. HMC is the main non-profit health care provider that manages ten highly specialized hospitals and PHCC that has 28 health centers. Testing is available to anyone with new continuous respiratory symptoms or anyone who is a contact of a person with a confirmed case. Tests done also include random samples tested for surveillance purposes, pre and post travel tests and individual test requests. Data on the *first positive test* for those tested were extracted from all tests conducted during 1 Jan 2021 up to 20 Feb 2022.

### Statistical analysis

Description statistics were used to describe the infected population. Cases and controls were matched in a 1:30 ratio on calendar month of infection and comorbidity category with an exact match used. Case participants were those requiring intensive care and controls were those requiring normal care.

Time interval in relation to the vaccination schedule was included as an independent variable, and effectiveness was assessed using a conditional logistic regression model. Vaccine effectiveness was defined as 1 minus adjusted odds ratio of requiring intensive care. The main confounders were age, calendar month of infection and comorbidity group, with the latter two being matched for. Vaccine effectiveness was adjusted in the conditional logistic regression model for age (continuous in years modeled using restricted cubic splines with four knots). Secondary analyses were not possible for type of vaccine (BNT162b2, or mRNA-1273 vaccine) as the majority received the BNT162b2 vaccine and data was sparse when thus stratified. Ethnicity of the person (Qatari or non-Qatari) or gender were not considered confounders as an independent association with time in relation to vaccination is unlikely given the equal access to health care for all residents of Qatar.

Age specific absolute risk reduction (ARR) was computed using effectiveness results from the conditional logistic regression and age-specific baseline risk (of requiring intensive care) estimated from the whole population and used to derive a second estimate of vaccine effectiveness - the number needed to vaccinate (NNV) to prevent one more case of COVID-19 requiring intensive care. This is computed as 1/ARR and provides a different perspective because the latter combines vaccine effectiveness with background risk of requiring intensive care. The main driver of background risk is patient age, the latter being the most critical determinant of the risk of requiring intensive care^11,12^.

No exclusions were made for the AstraZeneca vaccine as its frequency was too small to influence assessment of the mRNA vaccines, but a sensitivity analysis was carried out after exclusion of those individuals. Goodness of link was assessed via the linktest in Stata and goodness of fit of the model was assessed using McFadden’s R^2^ where 0.2 to 0.4 represent an excellent fit^13^. All analyses were conducted using Stata Version 15, College Station, TX, USA.

## RESULTS

### Descriptive Characteristics

Of the entire cohort of first infections reported between 1 Jan 2021 and 20 Feb 2022 in the State of Qatar, 98.99% had non-missing data needed for this study and 2113 of the latter progressed to require intensive care. Median interval between the first and second dose was 21 days (IQR 21 - 22) for the BNT162b2 vaccine and 28 days (IQR 28 – 30) for the mRNA-1273 vaccine. The median interval from the second to the third (booster) dose was 246 days (IQR 219 – 273). Of this population with first infections, 58% had this before vaccination, 36.8% during or after the primary dose schedule and 5.2% after the third dose. The median interval between the third dose and detection of the first infection, in those who tested positive, was 33 days (IQR 13 – 51). The predominant circulating variants in the wave in March-April 2021 were the B.1.1.7 (or alpha) and B.1.351 (or beta) variants^4^ while in the Dec 2021 – Jan 2022 wave was the B.1.1.529 (omicron) variant (Figure 1).

**Figure 1:**
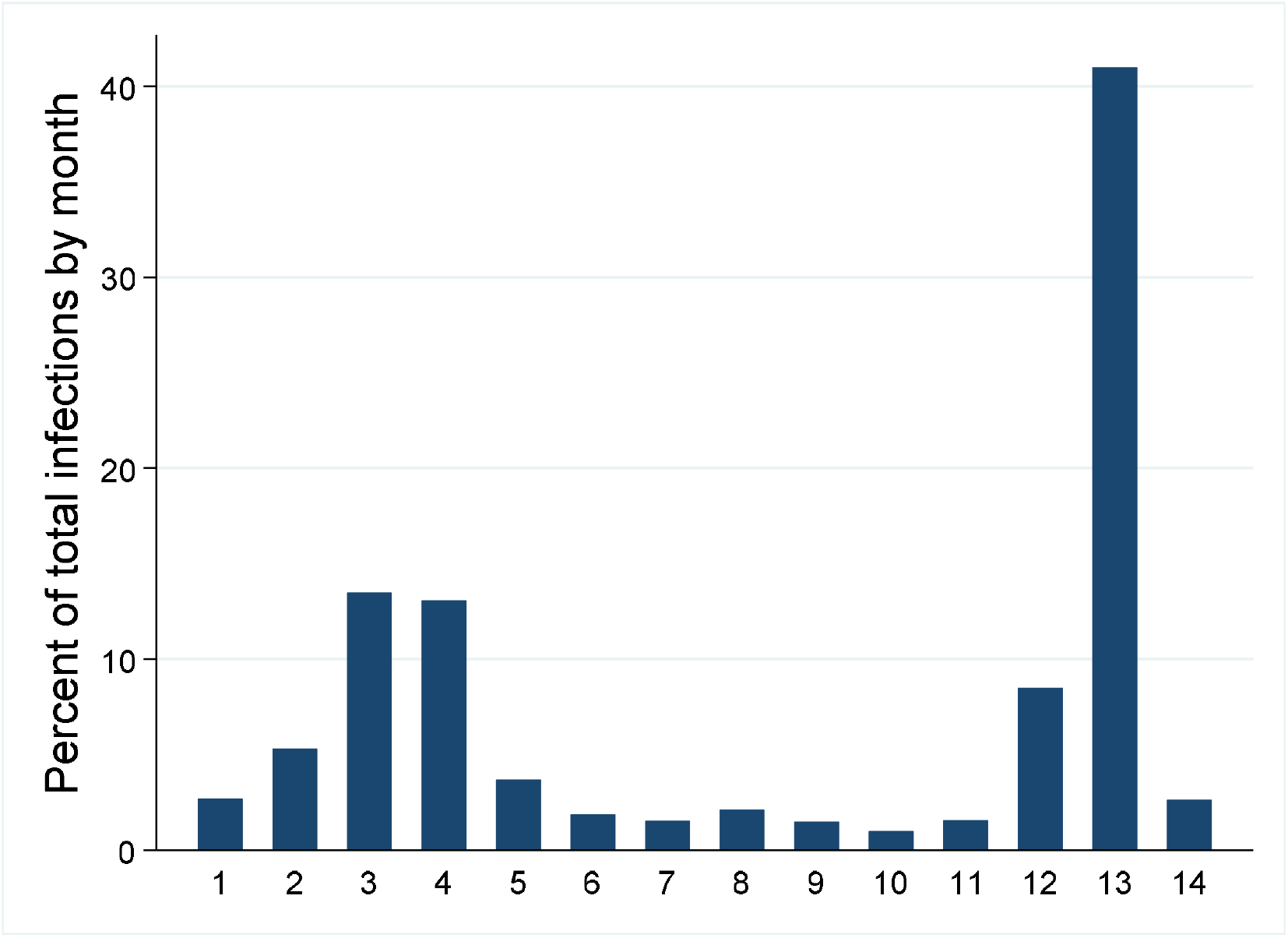
The two major waves are depicted over the 14 month study period (1-12 are Jan – Dec 2021 and 13-14 are Jan-Feb 2022).

After exact matching in a 1:30 ratio of cases and controls, 62,555 individuals were recruited into the matched case-control study. Matching (calendar month and comorbidity group) was not successful for 89 cases (4.1%) and the rest received 3 – 30 matched controls with 2089 (95.3%) having 30 matched controls. The distribution of the individuals in the matched case-control study (stratified by case and control status) in relation to age, sex, time interval (in relation to vaccination), comorbidity category, ethnic group and vaccine type is reported in Table 1.

**Table 1.**
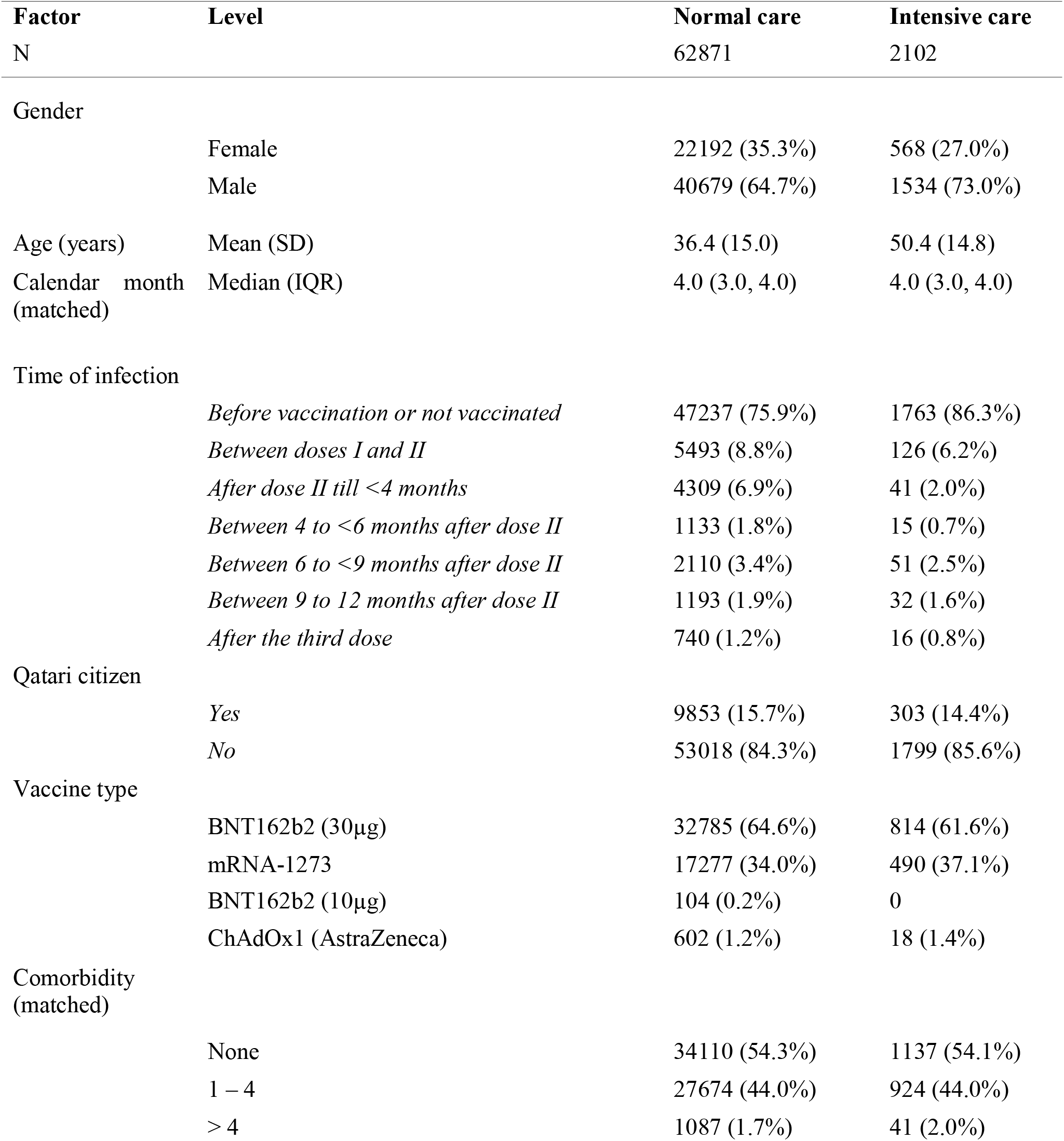
Baseline characteristics of the participants in the matched case control study

### Vaccine Effectiveness

Vaccine effectiveness against requiring intensive care in persons who received a primary course of the BNT162b2 or mRNA-1273 vaccine according to time interval in relation to primary immunization anchoring at the date of the second dose of the vaccines is reported in Figure 2. Vaccine effectiveness was 59% (95% confidence interval [CI], 50 to 76) between the first and second dose and strengthened to 89% (95% CI, 85 to 92) between the second dose and 4 months post the second dose. Vaccine effectiveness remained at this level (91%; 95% CI 84 to 95)) between 4 to 6 months after the second dose, at 6 to 9 months after the second dose (90%; 95% CI 84 to 94)) and at 9-12 months after the second dose (94%; 95% CI 89 to 97)). After the third dose (booster vaccine) effectiveness had strengthened to 95% (95% CI, 91 to 98). Goodness of link and fit of the regression model were both assessed to be satisfactory. There was no appreciable difference in vaccine effectiveness results when the case-control study was built and analyzed after exclusion of individuals who had taken the AstraZeneca vaccine.

**Figure 2.**
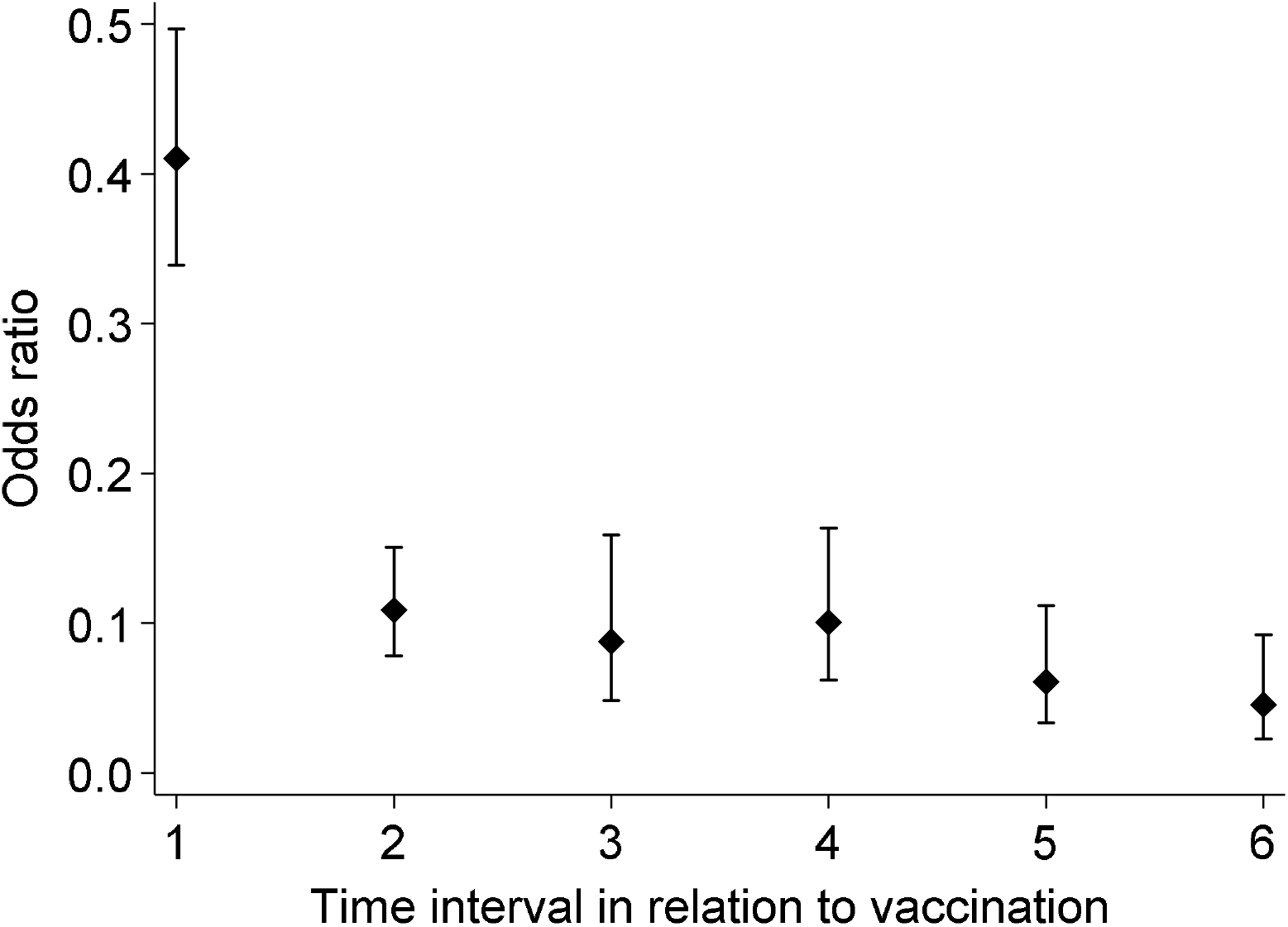
Odds ratios of requiring intensive care by interval in relation to vaccination from the matched case-control study *(Reference are those unvaccinated; 1=those infected between first and second dose; 2= those infected between the second dose to four months later; 3= those infected between the fourth to sixth month after the second dose; 4= those infected between the sixth to ninth month after the second dose; 5= those infected between the ninth to twelfth month after the second dose; 6= those infected after the third dose)*

As expected, risk of COVID-19 requiring intensive care in the study participants was dependent on the patients age and the NNV increased from 9 at age sixty (estimated unvaccinated risk of requiring intensive care 13%) to 90 (estimated unvaccinated risk of requiring intensive care 1.2%) at age thirty in the interval between nine and twelve months of the second dose.

## DISCUSSION

This study demonstrates that vaccine effectiveness for severe COVID-19 does not drop between 1 month and 12 months after the final vaccine dose. This contrasts with a recent meta-analysis ^9^ that included 12 studies evaluating vaccine efficacy or effectiveness over time for severe COVID-19 that reported that an average decrease by 10.0 percentage points (95% CI 6.1–15.4) among people of all ages and by 9.5 percentage points (5.7–14.6) among older people between 1 month and 6 months after the final vaccine dose. In this study, at 9 – 12 months after the second dose, vaccine effectiveness against requirement for intensive care remained high at 94%.

A larger decrease in the vaccine efficacy or effectiveness over time reported in the literature are not likely to be variant related ^9^ and therefore may be design related. The observational designs used previously include mainly test-negative design case-control studies and retrospective or prospective cohort studies and these are prone to unmeasured biases^9,14^ such as temporal trends for people who are vaccinated earlier being at sustained increased risk of infection compared with those who were vaccinated later; change in behavior after vaccination, temporal changes in testing frequency over time and differences in infection-derived immunity in the unvaccinated that may all lead to greater reductions in vaccine efficacy or effectiveness. This study avoided many of these biases by examining a complete cohort of COVID-19 cases in the State of Qatar in a defined period and then examining vaccine effectiveness in terms of progression to severity. Temporal differences were accounted for by matching on calendar month. In addition, comorbidities are difficult to model because of collinearity with age and were also matched by group. The key remaining confounder in this design was age, and this was dealt with robustly within the analysis using continuous age and restricted cubic splines for expected non-linearity. The handling of age has been a major pitfall of previous studies as matching on 10-year age-groups, for example, may result in significant residual confounding, and age is a very critical factor in progression to severe disease requiring intensive care.

The mRNA vaccines result in early production of serum IgA, IgM, and IgG antibodies^15,16^ and they have also been reported to be 80% effective for infection at least 14 days following the first dose ^17^. While vaccine effectiveness against SARS-CoV-2 infection tends to decline after 5 months^4^, protection against severe disease is sustained and we report here, that significant protection is also evident for severe disease in the interval between the two doses. The waning of protection against infection is consistent with studies that evaluated antibody titers after mRNA vaccination which demonstrate sustained titers of anti-spike and anti-receptor binding domain (RBD) IgG antibodies at 4 months after the second dose of the vaccine with a drop between 6-8 months. Interestingly the elevated antibody titers are likely linked to persistence of exosomal spike proteins at 4 months post vaccination. Studies are, however, limited by paucity of data beyond these timelines^18–23^. It is notable that most of the above mentioned studies failed to report the levels of neutralizing antibodies and T cell responses directed towards SARS-CoV-2 that are paramount in conferring longer term protection^1^ and studies have demonstrated that vaccination induces long-lasting memory B- and T-cell responses ^24–26^. This is consistent with the observation that natural infection with SARS-CoV-2 leads to a robust adaptive memory response that remains fairly constant 6-12 months post infection^27^. Also, most studies attempt to examine memory immune response to SARS-CoV-2 in peripheral blood from donors that typically lack the memory T and B cells repertoire while abundant SARS-CoV-2 reactive memory T and B cells reside in pulmonary lymph nodes with active germinal centers harboring SARS-CoV-2 specific follicular T helper cells that persist at least 6 months after resolution of infection^28^. The presence of T follicular helper cells in germinal centers indicates active affinity maturation with diverse antibody production conferring enhanced protection^28,29^. Indeed publications from our group^30,31^ and several others have shown a direct correlation of decreased lymphocyte count with COVID-19 severity and mortality while higher lymphocyte counts confer protection^32–34^. Interestingly, increased pro-inflammatory myeloid cells in the lung tissue and peripheral blood correlate with mortality and age ^35^. It is likely therefore that the sustained protection against requiring intensive care results from a sustained adaptive memory response.

To conclude, we demonstrate that effectiveness against requiring intensive care is sustained till at least 12 months after the second dose with no evidence of waning. We report here the NNV and not just relative odds reductions to avoid reporting biases that can then affect the interpretation of vaccine effectiveness for policy makers ^36^. The NNV is strongly age-dependent as we demonstrate here and therefore just looking at the relative summary measure for effectiveness fails to put the effectiveness results in context. In other words, the most vulnerable groups have lower NNVs given that their baseline risks of severe disease are larger. This study confirms that there is no waning of effectiveness against severe disease till 12 months and these data therefore support the conclusion that protection against severe infection is robust till at least 12 months but perhaps longer and a booster shot at 12 months can be a reasonable policy decision (although a longer interval could also be a possibility). Future studies should report age specific NNVs in addition to vaccine effectiveness or efficacy over time and extend follow-up beyond 12 months as these are the outcomes that will help us consolidate COVID-19 policy decisions.

## Data Availability

All data produced in the present work are summarized in the manuscript

## ACKNOWLEDGEMENTS

We thank our colleagues at the Ministry of Public Health-Qatar, Hamad Medical Corporation, Primary Health Care Corporation, and Qatar University for their earnest efforts and contributions that made this study possible.

